# Hospital admissions among adolescents with local authority care experience or special educational needs in England: a population-based cohort study using linked administrative data from health, education and social care services

**DOI:** 10.1101/2025.04.01.25325015

**Authors:** R Blackburn, T Kokosi, M Heys, R Gilbert, Zylbersztejn

**Affiliations:** UCL Great Ormond Street Institute of Child Health, 30 Guilford Street, London WC1N 1EH; NIHR Biomedical Research Centre, Great Ormond Street Hospital for Children NHS Foundation Trust, London, UK

## Abstract

**Background:** We aimed to quantify planned and unplanned hospital admissions during adolescence for pupils with special educational needs (SEN) and/or experience of care under local authority supervision.

**Methods:** We analysed linked hospital, education and social care records with whole nation coverage of England (ECHILD). The cohort comprised pupils who started secondary school between 2007/8 and 2011/12 (aged 11 years), with follow-up until March 2020 (until 18-23 years old). Hospital admission rates were examined by gender and age across distinct groups with varying levels of statutory support during school: i) no support, ii) school-based SEN support only, iii) an Education and Health Care Plan (EHCP) only, iv) care-experienced only, v) SEN support and care-experienced, and vi) EHCP and care-experienced. Rate ratios were estimated for each group relative to no support using negative binomial regression models.

**Results:** The cohort included 2,807,230 pupils; 64% (1,791,190) received no support, 31% (876,400) SEN support only, 3% (84,410) EHCP only, 1% (29,330) SEN support and care-experience, with the remaining groups accounting for <1%. Unplanned admissions were higher in all groups with support than peers with no support, and highest in care-experienced girls who had an EHCP (21.9 (95%CI; 21.4-22.4) per 100 person-years). Almost half of all unplanned admissions were mental health-related. Pregnancy-related admission rates were highest in care-experienced girls (14 per 100 person-years).

**Conclusions:** We found evidence of high levels of unplanned admissions coupled with low levels of planned care use for pupils with multiple needs, indicating a need for preventative care.

**KEY MESSAGES:**

- **What is already known on this topic** – previous research indicates a higher burden of physical and mental health needs among children and young people who are care-experienced and/or have additional educational needs. Although these young people are known to services, there are concerns about the accessibility and effectiveness of their care, which may be fragmented across sectors and between child and adult services.
- **What this study adds** – this research takes a multi-sectoral approach to quantify planned and unplanned hospital admissions during adolescence for children in England who are care-experienced and/or have additional educational needs. The study identified high levels of unplanned hospital use coupled with low levels of planned use for adolescents with multiple needs, thus indicating a need for earlier and preventative care.
- **How this study might affect research, practice or policy** – this research provides evidence of the multifaceted service needs of young people spanning physical, mental and reproductive health, and education and care systems, and highlights the potential benefits of greater co-ordination of early and effective care across services.

## INTRODUCTION

Children and young people (CYP) who receive statutory support from Children’s Social Care or Education services more frequently have additional long-term physical and mental health needs than their peers.(1–3) By age 18 years, 3-4% of CYP have ever been under local authority supervision (often with family or foster carers) and are termed Children Looked After (CLA).(4) These care-experienced CYP more frequently experience adverse childhood experiences such as physical, sexual or emotional abuse, and poorer long-term health, education and employment outcomes than peers who never experience out-of-home care.(5) Special Educational Needs (SEN) provision refers to additional support for children with health, learning or behavioural problems that impact their ability to learn. Over a third of CYP receive SEN provision during school ages,(6) including those with an education, health, and care plan (EHCP). An EHCP involves a formal assessment requested by the school or parents, followed by a plan of care and additional funding for support up to age 25 years. In contrast, SEN support (formerly School Action/Plus) is less formalised, often inconsistent, funded by a block notional budget to schools allocated based on a national formula,(7) and is given at the discretion of the school as part of the usual curriculum (e.g. extra help with communication or physical needs, special learning programmes). Social care and educational needs are often linked; by Year 11 (age 16) over 80% of pupils who ever experienced care have SEN recorded, and 23% have an EHCP.(6)

Adolescents experience multiple transitions and changes including ageing out of children’s services for health, education and social care that are structured to provide greater continuity and holistic care than adult services.(8) Service transitions are typically associated with marked rises in unplanned hospital admissions with physical and mental causes for CYP aged 16-18 years.(9) Health in adolescence is an important indicator of adult health, with three-quarters of life-time mental health conditions having onset by age 25 years.(10) Poor childhood mental health is a particular concern, with increased recognition that CYP community services are unable to meet demand. This impacts on the use of acute hospital care for CYP who are admitted to hospital (e.g., 19% of paediatric unplanned hospital admissions for 11-15 years olds were for mental health-related problems).(11) In CYP, manifestations of stress/distress can also present more broadly as physical symptoms (e.g. medically unexplained pain), or as behaviours including self-harm or substance misuse.(12)

Whilst it is known that CYP receiving support for social care or SEN have greater health needs, there is limited evidence quantifying how use of healthcare services changes during the adolescent period for these groups. Previous research suggests that CYP with SEN or who are care-experienced less frequently receive appropriate out-of-hospital care(1,8,13), likely contributing to poorer long-term physical and mental health.(5,14,15) Understanding patterns of health service demand within population groups, and over time, can help inform intervention points and service configuration.(8,16–18) We therefore aimed to quantify planned and unplanned hospital admissions for groups of CYP in England with anticipated differences in health, education and social care needs as indicated by records of care-experience and/or having SEN provision, within strata of age and gender. We also investigate the subset of unplanned hospital admissions indicative of mental health or related psychosocial needs because these admissions are extremely distressing for the young person, expensive and inefficient for health services, and may be preventable through earlier intervention and support in community settings.

## METHODS

### Data source

We used the Education and Child Health Insights from Linked Data (ECHILD) database of de-identified administrative health, education, and social care records for all CYP in England.(19,20) Hospital Episodes Statistics (HES) captures details of all NHS-funded hospital admissions,(21) including patient demographics and International Classification of Diseases (ICD10) coded diagnostic data. Information about pupil characteristics and enrolment in all state-funded education (93% of all pupils) is captured in termly school censuses from the National Pupil Database (NPD)(22), including linked information on CLA status.(23) Data availability changes over time, resulting in different follow-up durations for pupils of different ages (Figure 1).

**Figure 1.**
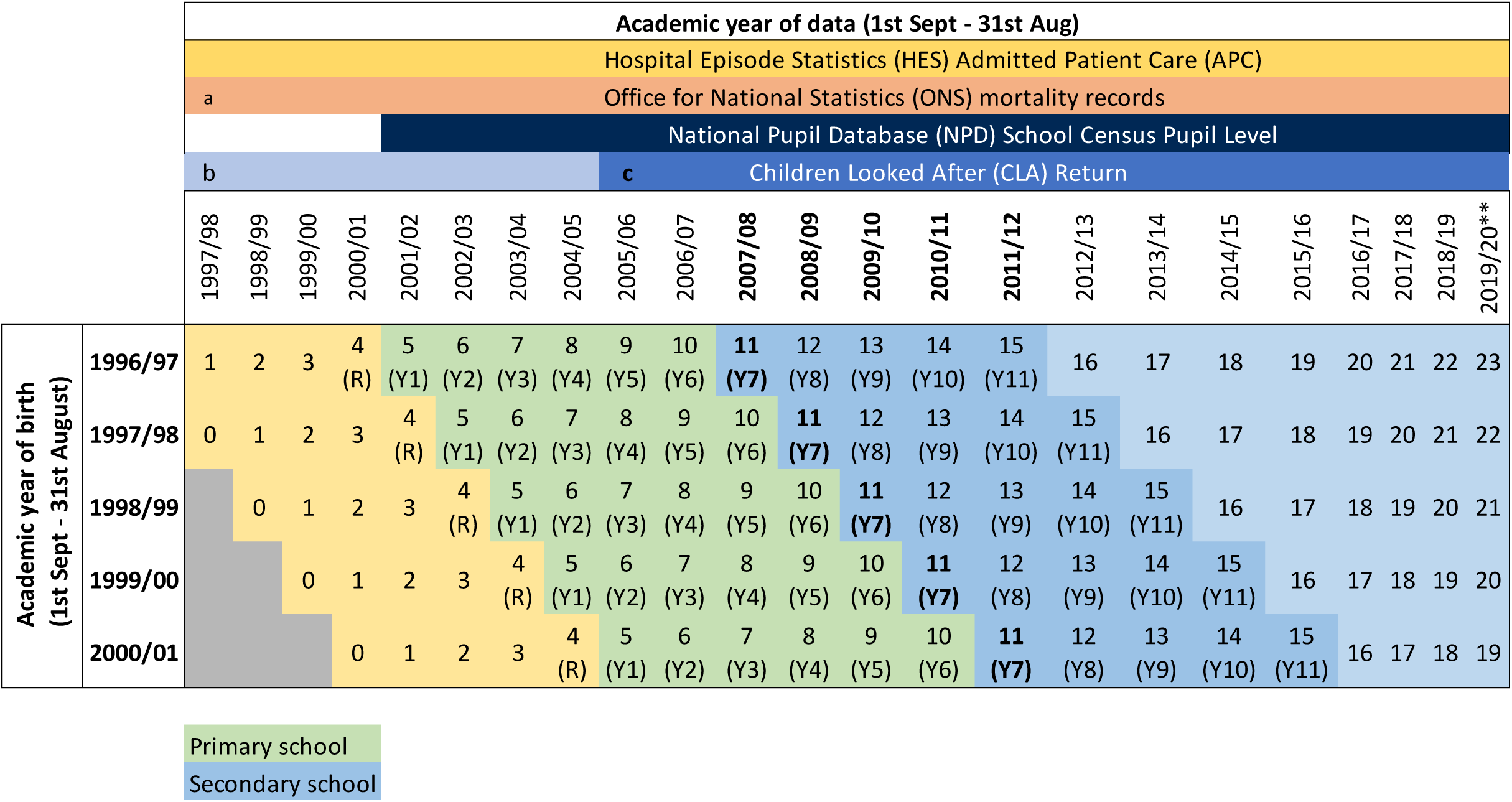
Cohort overview a: Partial coverage of an academic year as ONS Mortality data were first linked to HES in January 1998. b: Partial coverage of population as between 01/04/1992 & 31/03/2003, CLA data were only collected for a one-third sample (i.e., children with a day of birth divisible by 3). c: Partial coverage as linkage between NPD and CLA began on 1 April 2005 for CLA **follow-up until 1^st^ March 2020

### Study population

The study cohort included all pupils aged 11 years at the start of academic years 2007/08 to 2011/12 (born 1996/7-2000/01), who were enrolled in state-funded schools in England in Year 7 (or in specialist provision and not following the National Curriculum). We excluded pupils with unrecorded gender (*n*=34). CYP were followed up from 1^st^ September of Year 7 until 1^st^ March 2020, 24^th^ birthday or death, whichever occurred first. We were not able to account for out-of-country migration.

### Exposures

We indicated the highest-level of SEN provision (EHCPs including attending specialist provision, SEN support, none) ever recorded in any school census during primary school (Years 1-6). CLA records for children aged <5 years who have not started school are under-ascertained within ECHILD because the pseudo-identifiers for these records cannot be reliably linked to subsequent education records for same child. We therefore applied a broader time window for identifying care-experienced children, defined as a CLA record/s at any point before the end of secondary school (31^st^ August of the year they turn 16 years), to increase ascertainment and because experiences of adversity likely predate the first recorded care placement and may have enduring health impacts.

We derived 6 mutually exclusive groups of children with varying levels of statutory support from education and social care (no statutory support, SEN support only, EHCP only, CLA only, both EHCP and CLA, both SEN support and CLA) hypothesising that groups with greater complexity and intensity of support (e.g. EHCP vs SEN support) will also have additional healthcare demand.

### Characteristics and covariates

We extracted information on baseline (Spring Term, Year 7) pupil characteristics from NPD including month/year of birth, gender, free school meals eligibility (FMS), English as first language, Income Deprivation Affecting Children Index (IDACI) quintile of area-level deprivation and ethnic group (as defined in Table 1). We used ICD-10 codes recorded during hospital admissions from 1^st^ April 1997 until 1^st^ September of Year 7 to identify children with a chronic condition.(24)

**Table 1.** Overview of cohort characteristics.

|  | All children |  | Any SEN or CLA provision |  | Only EHCP |  | Only SEN support |  | Only CLA |  | EHCP and CLA |  | SEN Support & CLA |  |
| --- | --- | --- | --- | --- | --- | --- | --- | --- | --- | --- | --- | --- | --- | --- |
|  | N | % | N | % | N | % | N | % | N | % | N | % | N | % |
| Total | 2,807,230 |  | 1,016,040 | 36% | 84,410 | 3% | 876,400 | 31% | 13,260 | k | 12,660 | k | 29,330 | 1% |
| **Gender** |  |  |  |  |  |  |  |  |  |  |  |  |  |  |
| Females | 1,368,120 | 49% | 383,040 | 38% | 21,710 | 26% | 334,880 | 38% | 8,940 | 67% | 3,500 | 28% | 14,030 | 48% |
| Males | 1,439,110 | 51% | 633,000 | 62% | 62,700 | 74% | 541,520 | 62% | 4,320 | 33% | 9,160 | 72% | 15,300 | 52% |
| **Month of birth** |  |  |  |  |  |  |  |  |  |  |  |  |  |  |
| Sep-Oct | 481,040 | 17% | 141,150 | 14% | 13,000 | 15% | 118,570 | 14% | 2,710 | 20% | 2,180 | 17% | 4,690 | 16% |
| Nov-Dec | 459,590 | 16% | 148,040 | 15% | 13,330 | 16% | 125,430 | 14% | 2,410 | 18% | 2,150 | 17% | 4,720 | 16% |
| Jan-Feb | 451,920 | 16% | 157,260 | 15% | 13,510 | 16% | 134,490 | 15% | 2,410 | 18% | 2,070 | 16% | 4,780 | 16% |
| Mar-Apr | 461,480 | 16% | 170,640 | 17% | 14,140 | 17% | 147,540 | 17% | 2,070 | 16% | 2,110 | 17% | 4,790 | 16% |
| May-Jun | 469,570 | 17% | 188,230 | 19% | 14,860 | 18% | 164,360 | 19% | 1,930 | 15% | 2,080 | 16% | 5,000 | 17% |
| Jul-Aug | 483,620 | 17% | 210,720 | 21% | 15,570 | 18% | 186,010 | 21% | 1,730 | 13% | 2,080 | 16% | 5,340 | 18% |
| **Academic cohort** |  |  |  |  |  |  |  |  |  |  |  |  |  |  |
| 2007/8 | 565,810 | 20% | 202,780 | 20% | 16,850 | 20% | 175,820 | 20% | 2,410 | 18% | 2,460 | 19% | 5,260 | 18% |
| 2008/9 | 576,110 | 21% | 203,170 | 20% | 17,330 | 21% | 174,800 | 20% | 2,620 | 20% | 2,760 | 22% | 5,650 | 19% |
| 209/10 | 562,800 | 20% | 201,970 | 20% | 16,940 | 20% | 173,870 | 20% | 2,680 | 20% | 2,540 | 20% | 5,940 | 20% |
| 2010/11 | 557,940 | 20% | 204,840 | 20% | 16,870 | 20% | 176,490 | 20% | 2,800 | 21% | 2,530 | 20% | 6,150 | 21% |
| 2011/12 | 544,560 | 19% | 203,280 | 20% | 16,420 | 19% | 175,420 | 20% | 2,740 | 21% | 2,370 | 19% | 6,330 | 22% |
| **Ethnic group** |  |  |  |  |  |  |  |  |  |  |  |  |  |  |
| White | 2,258,520 | 80% | 807,420 | 79% | 68,960 | 82% | 695,230 | 79% | 9,460 | 71% | 10,600 | 84% | 23,160 | 79% |
| Asian | 229,400 | 8% | 80,540 | 8% | 5,680 | 7% | 72,410 | 8% | 990 | 7% | 400 | 3% | 1,050 | 4% |
| Black | 127,840 | 5% | 57,000 | 6% | 4,120 | 5% | 48,810 | 6% | 1,250 | 9% | 620 | 5% | 2,210 | 8% |
| Mixed | 108,230 | 4% | 40,600 | 4% | 3,070 | 4% | 33,770 | 4% | 1,030 | 8% | 650 | 5% | 2,080 | 7% |
| Other | 43,290 | 2% | 14,540 | 1% | 920 | 1% | 13,000 | 1% | 270 | 2% | 100 | 1% | 260 | 1% |
| Missing | 39,960 | 1% | 15,950 | 2% | 1,680 | 2% | 13,190 | 2% | 250 | 2% | 280 | 2% | 560 | 2% |
| **First language** |  |  |  |  |  |  |  |  |  |  |  |  |  |  |
| English | 2452530 | 87% | 881940 | 87% | 74060 | 88% | 758650 | 87% | 11160 | 84% | 11180 | 88% | 26900 | 92% |
| Other | 337410 | 12% | 125370 | 12% | 7750 | 9% | 112980 | 13% | 1980 | 15% | 550 | 4% | 2130 | 7% |
| Missing | 17280 | 1% | 8730 | 1% | 2600 | 3% | 4770 | 1% | 120 | 1% | 940 | 7% | 300 | 1% |
| **Free School meal eligibility** |  |  |  |  |  |  |  |  |  |  |  |  |  |  |
| No | 2,319,150 | 83% | 740,170 | 73% | 59,180 | 70% | 648,730 | 74% | 7,900 | 60% | 8,380 | 66% | 15,960 | 54% |
| Yes | 488,050 | 17% | 275,850 | 27% | 25,220 | 30% | 227,660 | 26% | 5,350 | 40% | 4,270 | 34% | 13,360 | 46% |
| Missing | 40 | k | 30 | k | 10 | k | 10 | k | c |  | c |  | c |  |
| **IDACI quintile** |  |  |  |  |  |  |  |  |  |  |  |  |  |  |
| Q1: most deprived | 669,230 | 24% | 318,550 | 31% | 24,170 | 29% | 274,720 | 31% | 4,950 | 37% | 3,310 | 26% | 11,410 | 39% |
| Q2 | 565,980 | 20% | 231,480 | 23% | 18,670 | 22% | 199,590 | 23% | 3,310 | 25% | 2,700 | 21% | 7,200 | 25% |
| Q3 | 528,330 | 19% | 181,610 | 18% | 14,880 | 18% | 157,520 | 18% | 2,210 | 17% | 2,240 | 18% | 4,770 | 16% |
| Q4 | 520,120 | 19% | 151,740 | 15% | 12,830 | 15% | 132,090 | 15% | 1,580 | 12% | 1,850 | 15% | 3,380 | 12% |
| Q5: least deprived | 510,840 | 18% | 125,690 | 12% | 11,240 | 13% | 109,550 | 13% | 1,100 | 8% | 1,550 | 12% | 2,250 | 8% |
| Missing | 12,740 | k | 6,970 | 1% | 2,620 | 3% | 2,930 | k | 100 | 1% | 1,010 | 8% | 320 | 1% |
| **Chronic conditions** |  |  |  |  |  |  |  |  |  |  |  |  |  |  |
| No | 2,383,760 | 85% | 810,510 | 80% | 48,280 | 57% | 720,740 | 82% | 11,470 | 87% | 6,200 | 49% | 23,820 | 81% |
| yes | 423,470 | 15% | 205,530 | 20% | 36,130 | 43% | 155,650 | 18% | 1,780 | 13% | 6,460 | 51% | 5,500 | 19% |
| **Died** | 5,800 | k | 3,390 | k | 880 | 1% | 1,970 | k | 80 | 1% | 310 | 2% | 150 | 1% |

CLA: child looked after; EHCP: education, health and care plan; IDACI; Income Deprivation Affecting Children Index; SEN: Special Educational Needs;
Data are presented in accordance with statistical control policy for linked CLA data such that: national figures are rounded to the nearest 10 and percentages are rounded to 0 decimal places 'c' in this table means the figures have been suppressed to protect confidentiality 'k' is used when a result that is not zero would appear as zero due to rounding

### Outcomes

Primary outcomes were planned and unplanned hospital admission rates.

Hospital admissions were defined as continuous time under care of NHS, therefore consecutive admissions within 1 day of discharge (e.g. hospital transfers) were treated as part of the same admission. We defined admission type (planned/unplanned) according to the admission method recorded at the start of admission.

Admissions related to pregnancy(9) were expected to increase with age and were excluded from the primary outcomes but are reported as a secondary outcome for girls. Information on pregnancy (particularly early pregnancy and loss) is poorly captured in HES, such that these admissions primarily reflect deliveries at 24 weeks gestation or more.

Unplanned mental health-related admissions were investigated as a secondary outcome and reflect diagnostic codes for:

– established and/or enduring mental health diagnoses defined using F chapter ICD-10 codes (mental, behavioural, and neurodevelopmental disorders) recorded as any diagnosis
– injuries relating to drug and alcohol misuse, maltreatment, violence, and intentional self-harm (including self-poisoning and self-cutting), recorded as any diagnosis.
– Potential manifestations of stress indicative of undifferentiated and/or transient symptoms recorded as the primary diagnosis, including medically unexplained abdominal pain. Specific medical exclusions (e.g. appendectomy) recorded as part of the same continuous inpatient stay were applied.

Codes are outlined in ECHILD Code Repository: https://code.echild.ac.uk/srp_nichobhthaigh_v2

### Statistical analysis

We describe numbers and proportions of CYP overall, by pupil characteristic and across statutory support groups. To comply with statistical disclosure rules all counts are rounded to the nearest 10, thus counts may not sum to the total study population.

We calculated hospital admission rates (planned, unplanned and unplanned mental health-related admissions) per 100 person years (100PY) at risk, which was calculated as the time from 1^st^ September of Year 7 until end of follow-up, excluding admitted time in hospital (including pregnancy-related admissions). Point estimates are reported with associated 95% confidence intervals (CI).

Crude hospital admissions rate ratios (RR) and 95% CI were estimated for pupils with different levels of statutory support compared to peers with no support using negative binomial regression (due to overdispersion). Models were stratified by gender and age (categorised as 11-12, 13-15, 16-17,18-23) and adjusted for year of birth to account for potential cohort effects. Data were analysed with R Studio and Stata version 18.(25,26)

## RESULTS

### Cohort characteristics

The study cohort included 1,368,120 girls (49%) and 1,439,110 (51%) boys. Overall, 36% (*n*=1,016,040; 62% were boys) of pupils received any statutory support (Appendix; Table 1): 31% (*n*=876,400; 62% boys) had SEN support only, 3% (*n*=84,410; 74% boys) had EHCP only, and <1% were care-experienced only (*n*=13,260, 33% boys), had an EHCP and were care-experienced (*n*=12,660, 72% boys), or SEN support and care-experience (*n*=29,330, 52% boys, Table 1). Of pupils who were care-experienced, 53% had recorded SEN support and 23% had an EHCP (76% any recorded SEN provision).

Pupils receiving any statutory services were more likely to be; eligible for free school meals (particularly care-experienced pupils), living in the most deprived 20% of areas, and have chronic conditions (particularly EHCP groups) compared to their peers. Between 1-2% of pupils who had an EHCP and/or were care-experienced died during the study period compared to <1% of peers with no support (Table 1).

### Hospital admission rates

#### Planned admission rates

More than 1 in 5 of all pupils had at least one planned admission, with significant variation by age and level of support (Appendix; Figure 1A/Table 3). Girls had higher rates of planned admissions than boys overall ((4.83/100PY (95% CI; 4.82-4.83) vs 4.50/100PY (4.49-4.50)), across all ages and exposure/support groups (Figure 2A). Planned admission rates were consistently higher in groups receiving any kind of statutory support than peers with no support, except for care-experienced boys without an EHCP/SEN (Figure 2A and 3A). For these groups of care-experienced boys, planned admission rates were 18-19% lower at age 13-15 years (RR; 0.82 (0.79-0.86)) and 18+ years (RR; 0.81 (0.78-0.84)) than for peers of the same age with no statutory support. Rates were highest for pupils of secondary school ages with EHCPs, especially 11-12 year-old girls (RR; 14.1 (13.8-14.5)) and boys (RR; 10.3 (10.2-10.5)) who were also care-experienced (Figure 2A and 3A). Relative differences in planned admission rates for children with an EHCP compared to peers with no support reduced with age (Figure 3A). This reflects a rapid decline in planned admissions among EHCP groups after secondary schools ages, coinciding with transition into adult healthcare services (Figure 2A). Conversely, among pupils with no support, and for pupils in SEN/CLA/both groups, planned admission rates increased with increasing age (Figure 2A).

**Figure 2.**
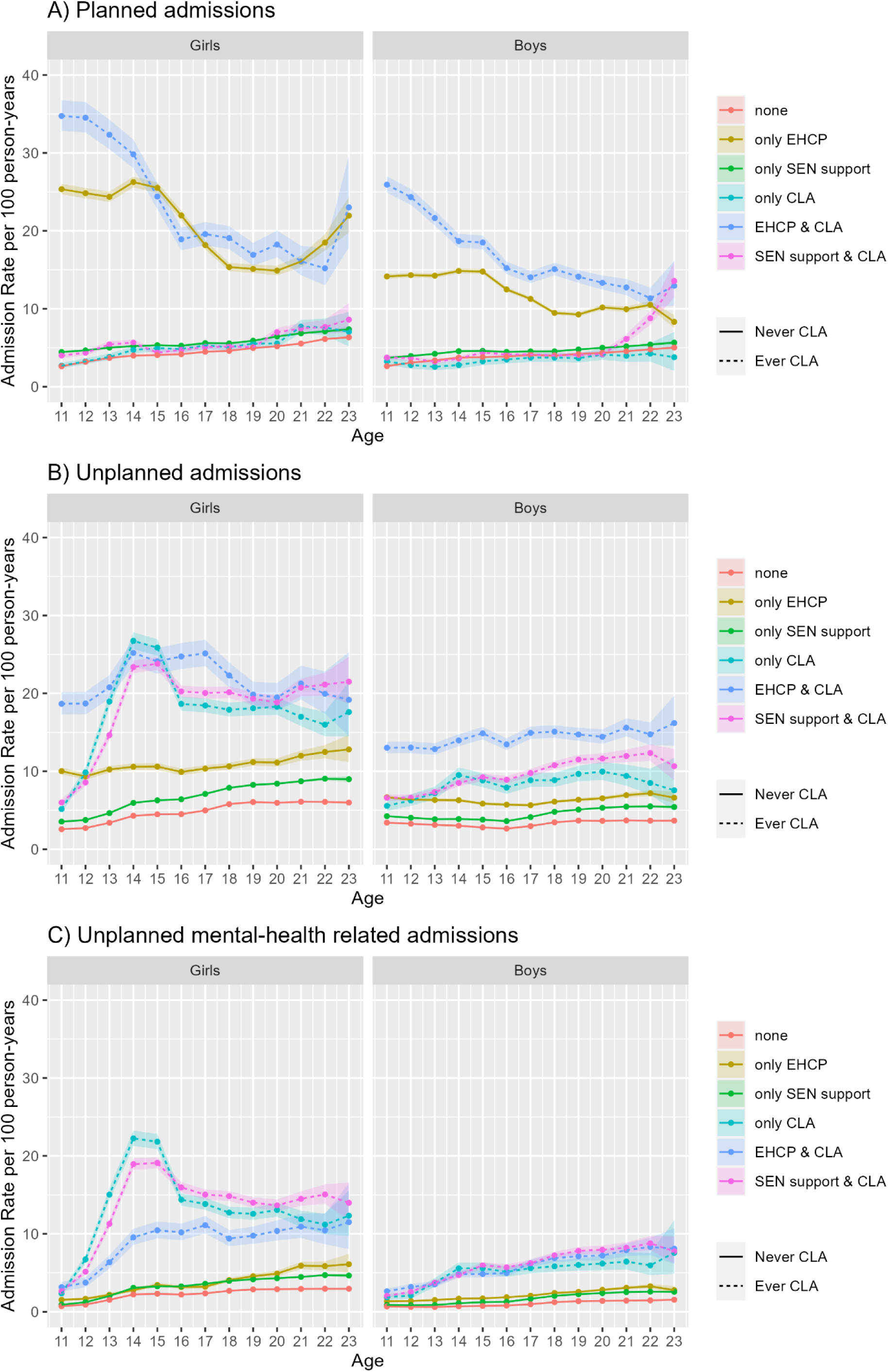
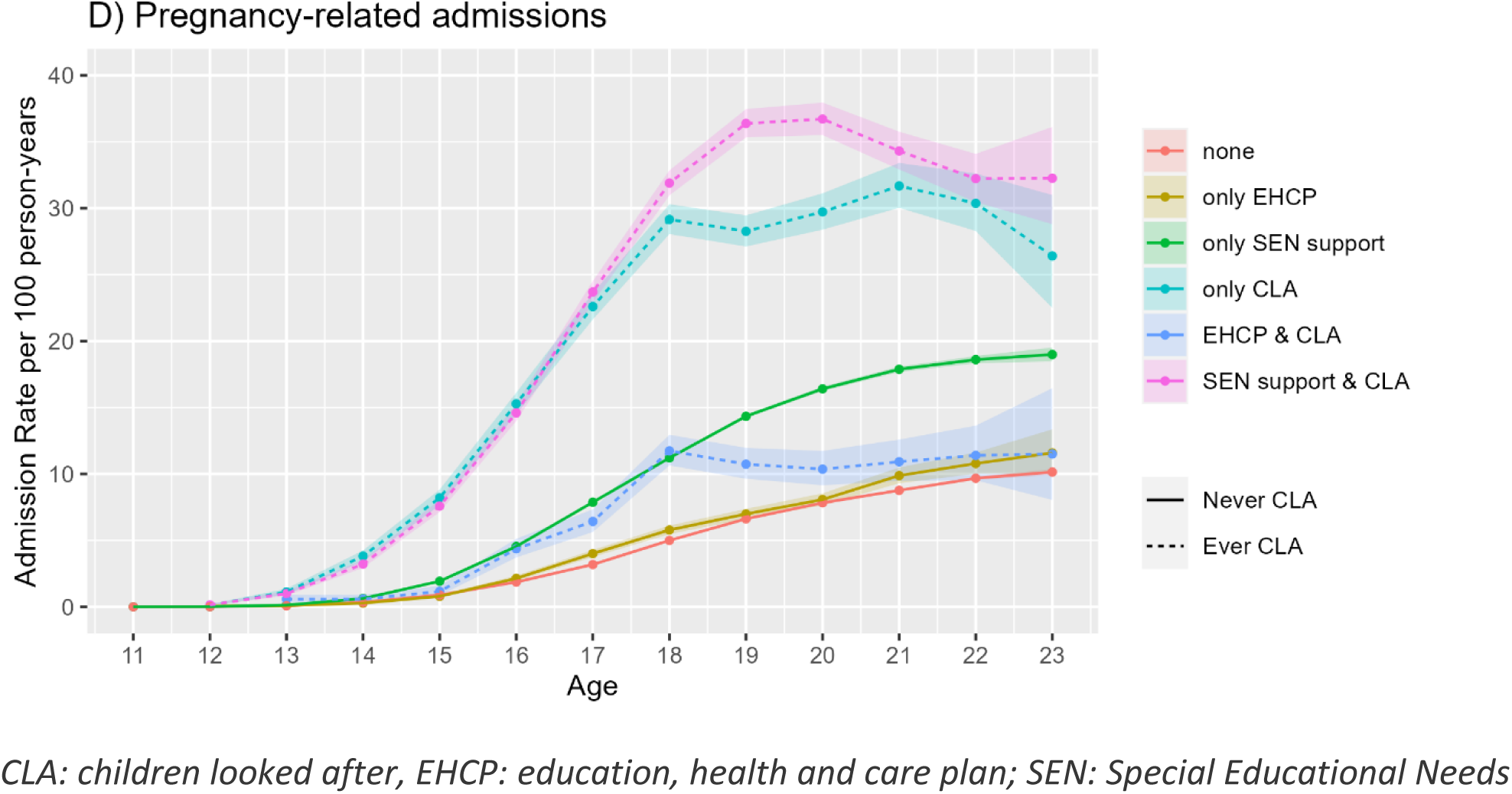
Hospital admission rates by age and exposure

**Figure 3.**
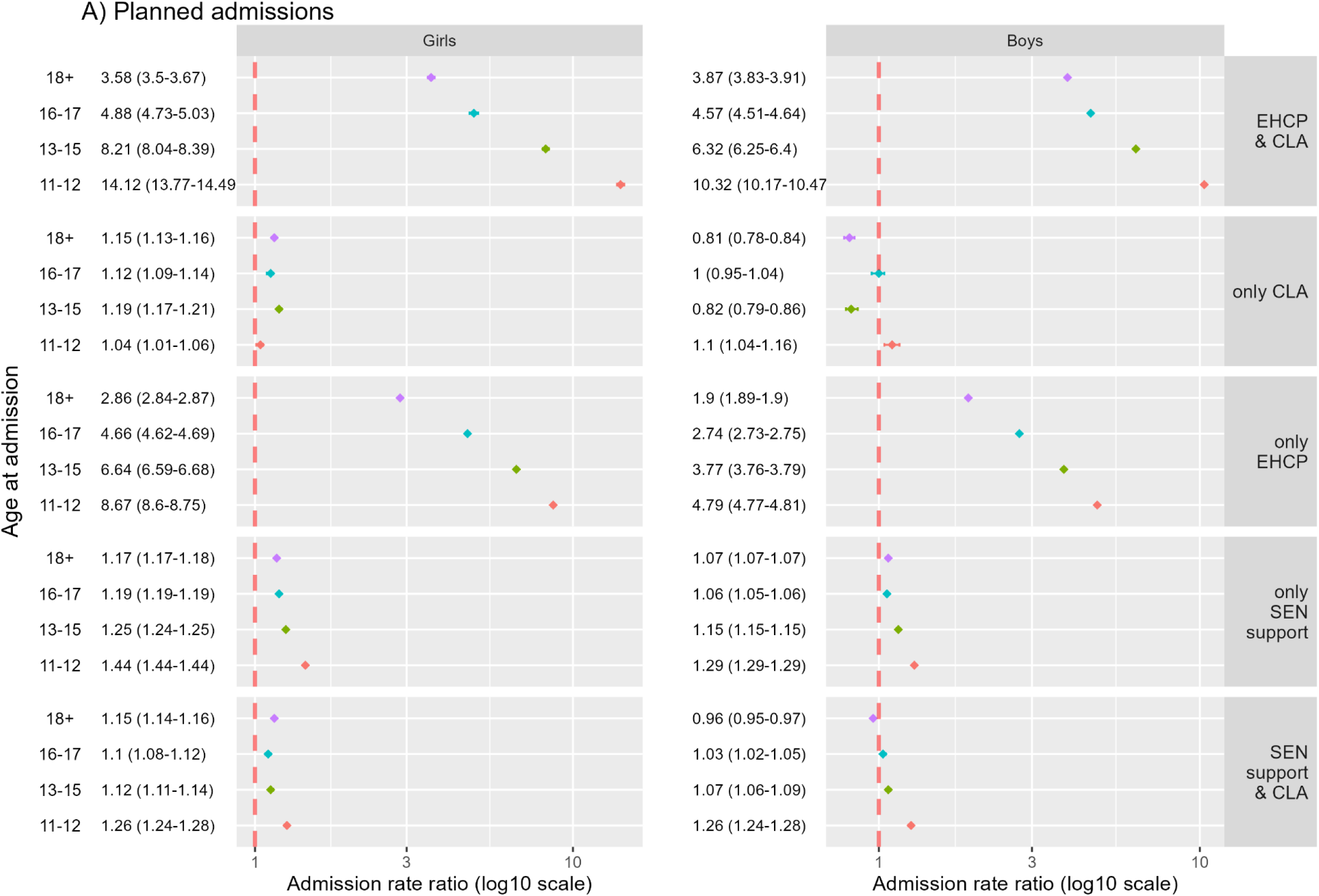
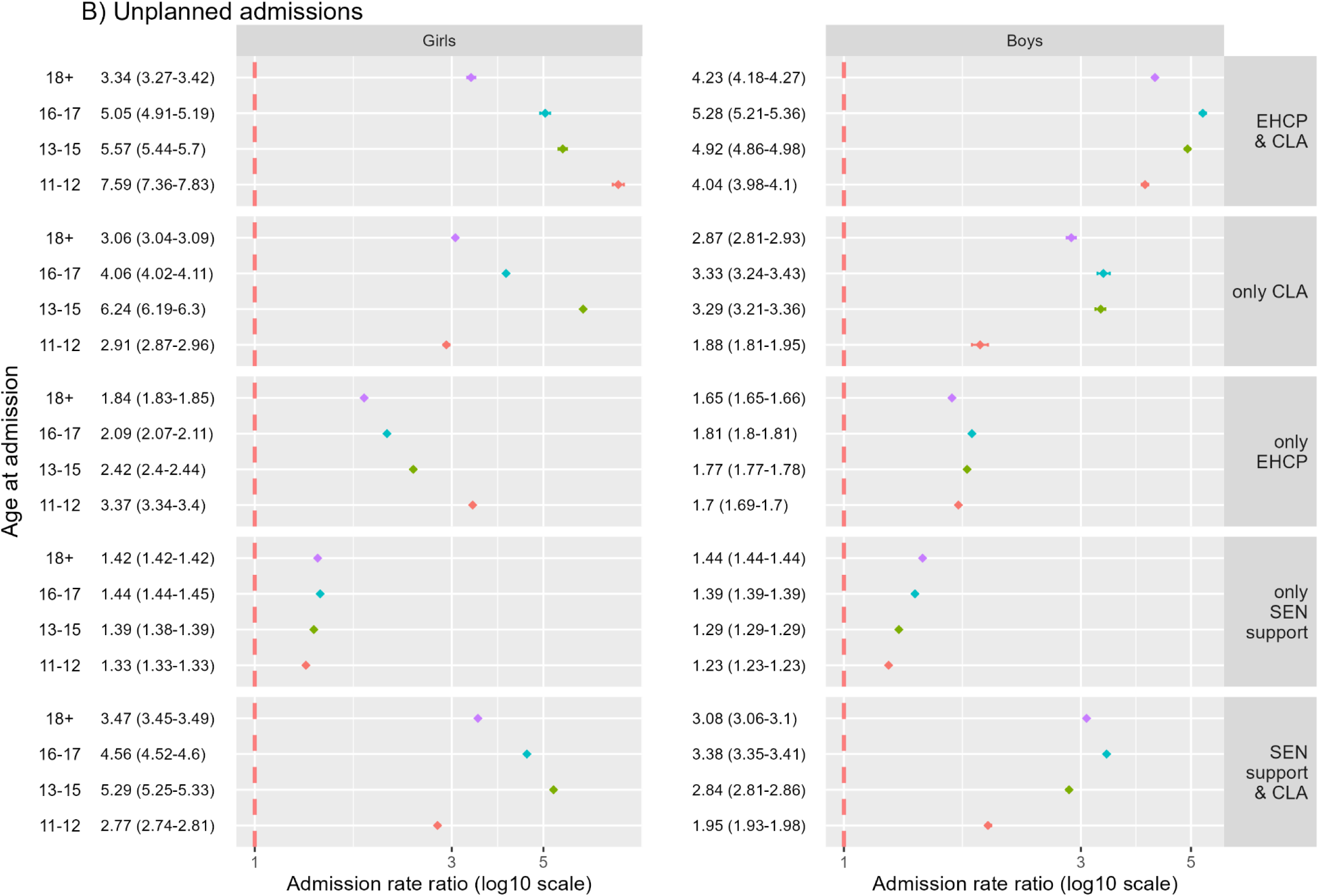
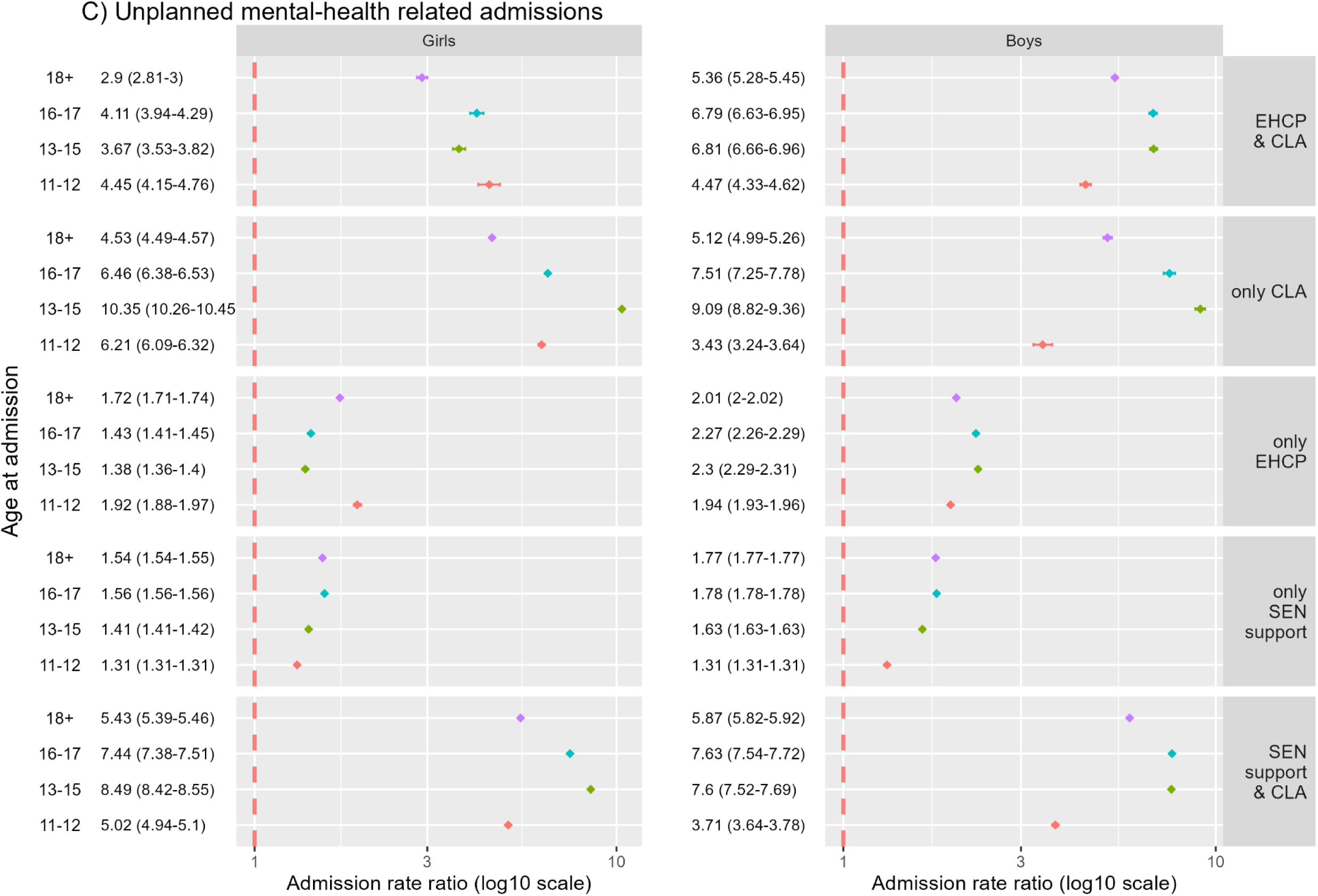
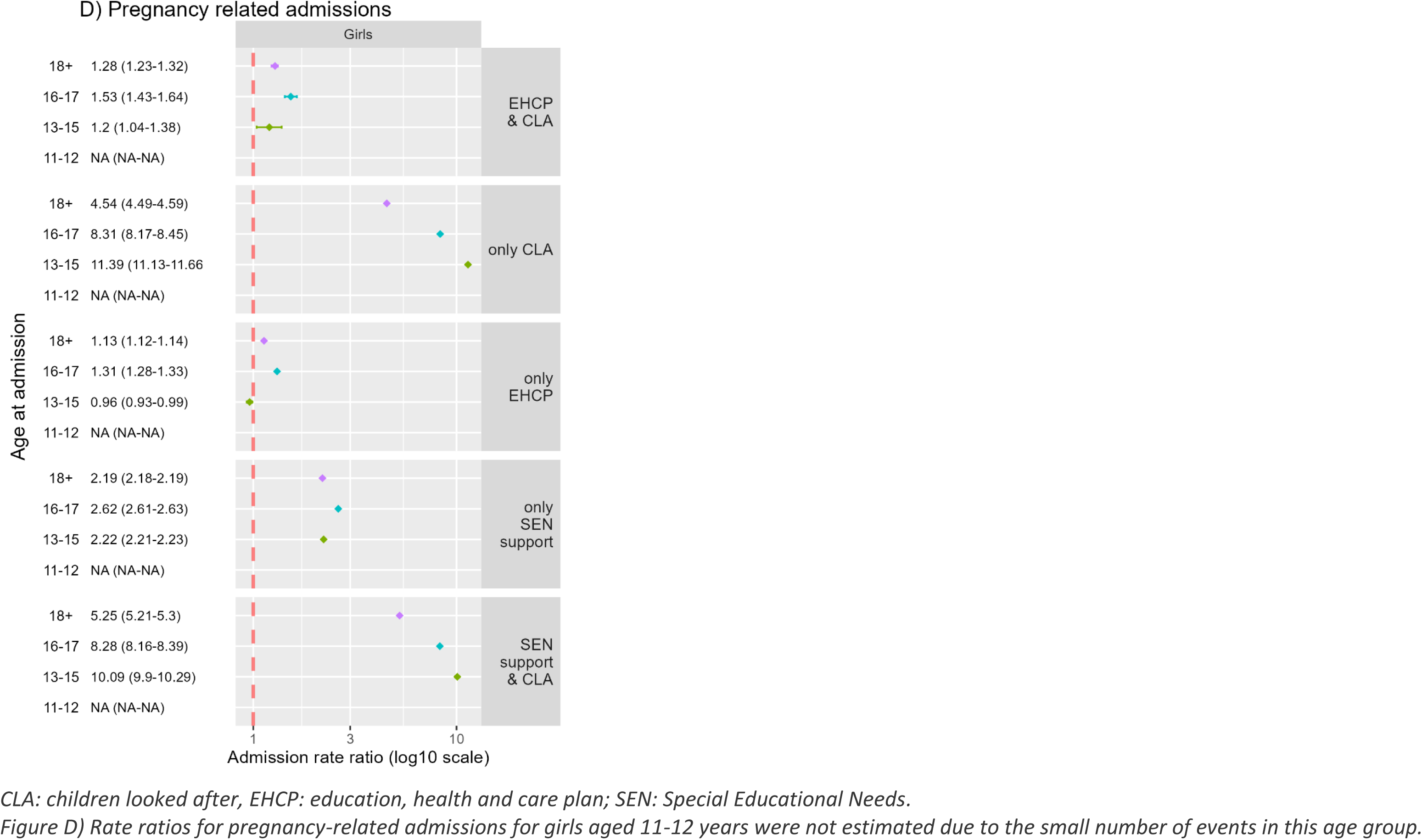
Rate ratios for planned (A), unplanned (B), unplanned mental-health related (C) and pregnancy-related (D) admissions by age and exposure group. Pupils with no support are the baseline group for all rate ratios.

#### Unplanned admission rates

One in four boys and girls had at least one unplanned hospital admission, with significant variation by age and level of support (Appendix; Figure 1B/Table 3). Girls had higher overall unplanned admission rates than boys (5.35/100PY (5.34-5.36) vs 3.92/100PY (3.91-3.93)) (Figure 2B).

Unplanned admission rates were higher among all groups with statutory support than peers with no support (Figures 2B and 3B; all RRs have 95%CI >1). Similar, but higher trends in unplanned admission rates were observed for pupils receiving SEN support only versus those with no support, and these differences widened with age (Figure 3B). Patterning of admission rates by age was particularly striking for girls (with a strong peak aged 14-15 years; Figure 2B), and especially for care-experienced groups where rates were over 5 times higher relative to peers with no support (Figure 3B, RR for girls aged 13-15 years: care-experienced only; 6.24 (6.19-6.30), EHCP and care-experience; 5.57 (5.44-5.7)), SEN and care-experience; 5.29 (5.25-5.33)). Care-experienced boys had 2-5 times higher rates of unplanned admissions than peers with no support and these differences were largest at age 16-17 years (Figure 3B, RR for boys aged 16-17 years: care-experienced only; 3.33 (3.24-3.42), EHCP and care-experience; 5.28 (5.21-5.36)), SEN and care-experience; 3.38 (3.35-3.41)).

#### Mental health-related admissions

Overall, 14% of girls and 9% of boys had at least one mental health-related admission, with significant variation by age and group (Appendix; Figure 1C/Table 3). Girls had higher overall rates of mental health-related admissions than boys (2.56/100PY (2.55-2,57) vs 1.28/100PY (1.28-1.29); Figure 2B). Overall, 48% of all unplanned admissions for girls and 33% for boys were mental health-related. Mental health-related unplanned admission rates were highest for care-experienced girls (12.6/100PY (12.47-12.74)) and boys (5.34/100PY (5.26-5.42))), accounting for 69% and 50% of all unplanned admissions, respectively. This compares to 2.11/100PY (2.10-2.12) for girls and 0.94/100PY (0.93-0.94) boys without any support, and accounting for 46% and 29% of all unplanned admissions, respectively (Appendix Table 2). For care-experienced girls, patterns of unplanned mental health-related admissions were similar to all unplanned admissions, peaking around age 14-15 years old. For boys, mental health-related admissions consistently increased with age (Figure 2C).

#### Pregnancy-related admissions

Overall, 14% of girls (4/100PY (3.99-4.01); Appendix; Tables 2/3) had at least one pregnancy-related admission: rates increased with age (Figure 2D/3D). Among care-experienced girls 39% (14/100PY (13.9-14.2)) had a pregnancy-related admission. Compared to girls with no support, pregnancy-related admission rates were raised in all care-experienced groups and were highest among 13–15-year-olds without an EHCP (RR aged 13-15 years: care-experienced only; 11.4 (11.1-11.7), SEN and care-experience; 10.1 (9.9-10.3); Figure 3D). Rates were twofold higher (Figure 3D) for girls with SEN support (only) compared to peers with no support (6.74/100PY (6.71-6.77)) vs 2.96/100PY (2.95-2.97)).

## DISCUSSION

This study takes a multi-sectoral approach to quantify the first whole nation estimates of hospital admissions for pupils in England receiving statutory support. Overall, more than 1 in 5 CYP in England experienced planned and unplanned hospital admissions, and unplanned admission rates were higher among all pupil groups with statutory support (versus pupils with no support). In contrast, rates of planned admissions were substantially higher for pupils with an EHCP (reflecting medical criteria for EHCP eligibility), but not in other groups receiving support suggesting potential unmet need.

Unplanned admissions were particularly high for care-experienced pupils, especially for those with coexisting SEN provision. Among care-experienced pupils, high proportions (half for boys, two thirds for girls) of unplanned admissions were mental health-related, indicating urgent unmet mental health needs. Rates of planned hospital admissions for care-experienced groups and no support groups were similar, except for care-experienced boys aged 13–15 and 18+ years where rates were one fifth lower. This is important because care-experienced pupils had higher rates of unplanned admissions, particularly for mental health. For girls, pregnancy-related admissions were higher for all groups receiving support, except for EHCP only. Over 1 in 3 care-experienced girls (versus 1 in 9 girls with no support) had a pregnancy-related admission during adolescence and rates were over tenfold higher among 13–15-year-olds, highlighting the sexual vulnerability of this group.

This study differentiates between groups of pupils receiving SEN and/or social care support to reflect differences in medical and social factors, e.g. greater medical needs among pupils with an EHCP, greater social adversity among care-experienced CYP. We report higher unplanned admission rates among girls (peaking at age 14-15 years), which mirrors other studies and may reflect biopsychosocial and structural factors (e.g. prioritising admissions for under 16s, particularly suspected self-harm) that warrants further mixed methods research.(11,12,27)

Our results suggest that care-experienced pupils, particularly boys, have similar or lower rates of planned care than pupils with no support. Analyses for Scotland report a higher prevalence of chronic health conditions and mental health admissions for care-experienced CYP than the general population,(28) indicating that our findings likely reflect unmet preventative care needs. Similarly raised unplanned admission rates have been reported for CYP in Wales with social care contact.(29). Care-experienced CYP often experience additional barriers to their peers, including inconsistent carer-giver support for healthcare seeking and access, and face particular challenges accessing mental health care.(29,30) High rates of mental health-related admissions to acute hospital wards do not reflect best practice for managing mental health and indicate that community and specialist services are not meeting demand.(29–31) We report high rates of pregnancy-related admissions for care-experienced girls, mirroring findings for Scotland, and raising concerns over sexual exploitation and safeguarding, as well as a need for intensive interventions such as the Family Nurse Partnership to limit intergenerational transmission of disadvantage.(3,32,33)

Strengths of the study include using longitudinal, multisector linked administrative datasets to examination health inequalities across statutory support groups, and whole-country coverage to minimise selection bias.(34) These groupings reflect pupils known to child health services, with actionable messages for social care and health. Limitations reflect variation in real-world testing and support for SEN and/or care-experience, such that need is imperfectly captured, and actual provision is unmeasured. CYP not enrolled in state-funded education for reasons reflecting school or child/family circumstances (e.g. pushing-out, independent education, home schooling) are an important group, have needs that are likely under-ascertained in this study and should be the focus of future research.(35) We applied a broader case definition (reflecting stress/distress-related presentations) that captures higher proportions of mental health-related admissions than other studies (e.g. 28% of admissions in 2021/22 for girls aged 11-15 years (11)). Further investigation of pathways across services and with additional linkage with adult service level data is needed, including NHS Child and Adolescent Mental Health Services,(36) which was not available for this study. Finally, our study does not reflect the pandemic or post-pandemic periods, and outcomes may have worsened following the erosion of youth services, with disproportionate impacts for CYP in care (37,38).

Our findings emphasise both greater healthcare needs for CYP with additional educational and/or social care needs (relative to peers) and imbalances in planned and unplanned hospital admissions. Action is most urgently needed for care-experienced CYP, those who are in mid-teens or aging out of children’s services. For these groups, rates of mental health-related admissions and teenage pregnancy are much higher than their peers, indicating that these young people do not receive appropriate support. Continued tailored support into post-16 education and social care must be young person-centred, trauma-informed and integrated across services. Finally, our results indicate a need for enhanced co-ordination of services spanning social care, education and health for young people with additional needs to ensure that care is seamless and effective.

## SUPPLEMENTARY MATERIALS

**Appendix Figure 1.**
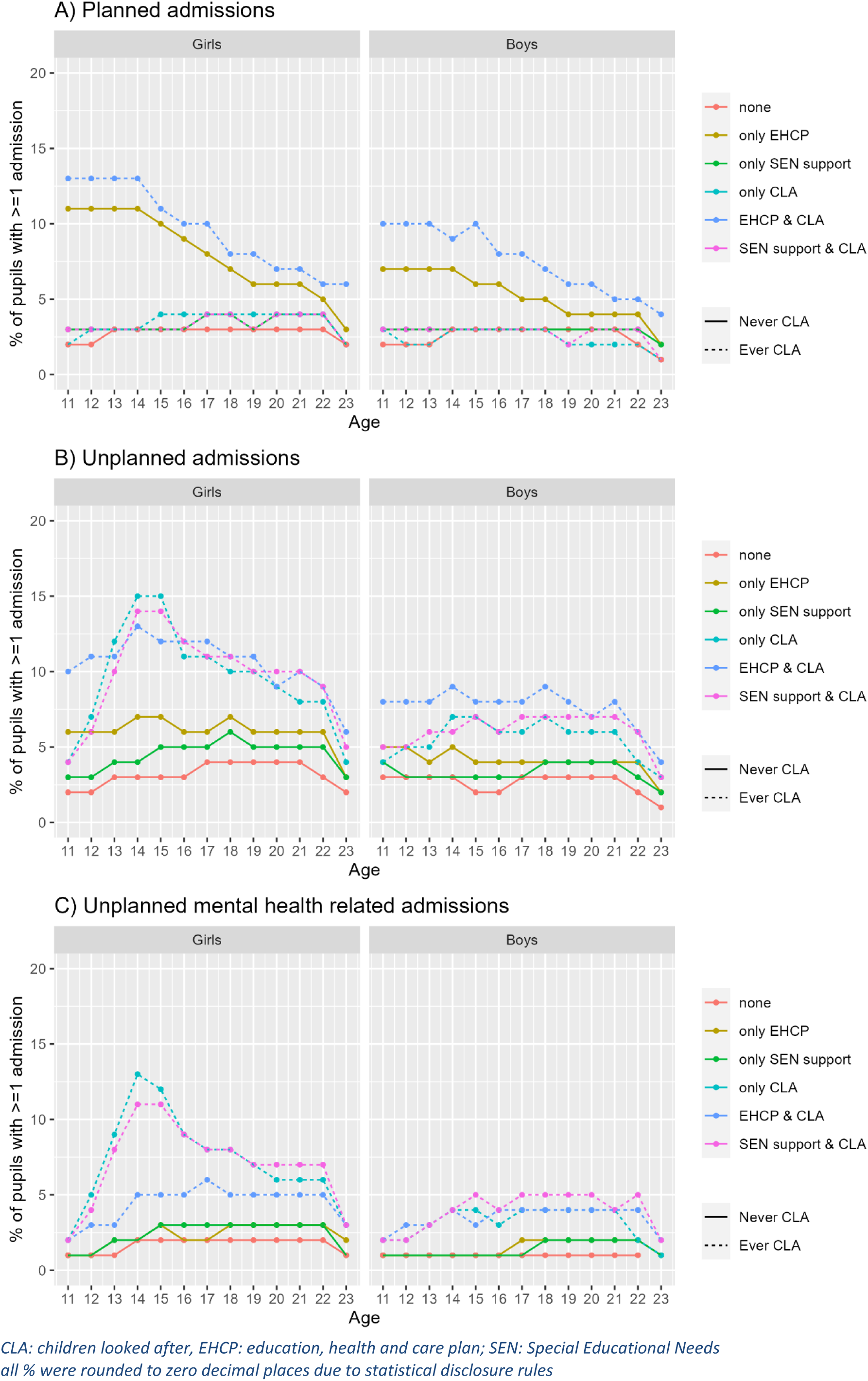
Proportion of children with at least one planned/unplanned admission by age and exposure group

**Appendix Table 1.**
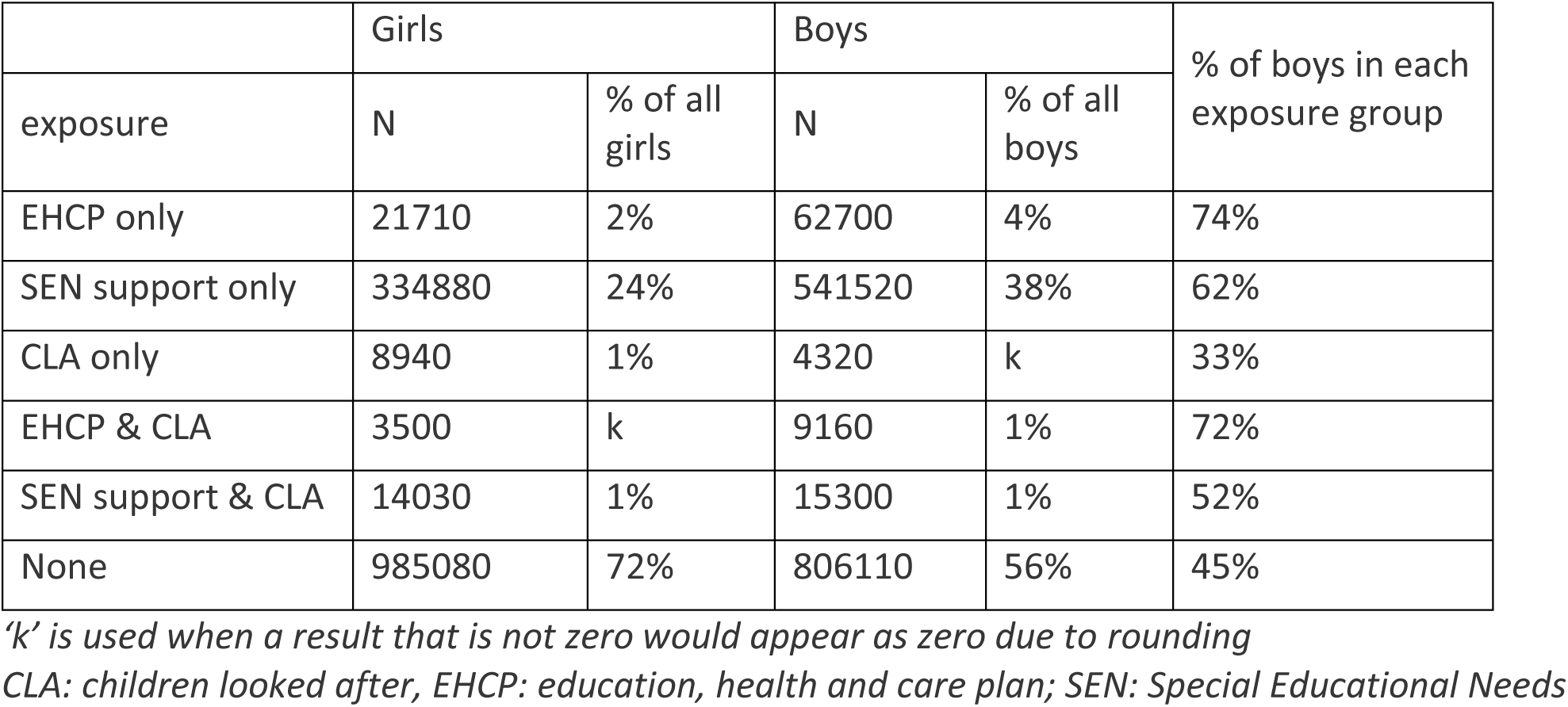
distribution of boys and girls in each exposure group.

| exposure | Girls |  | Boys |  | % of boys in each exposure group |
| --- | --- | --- | --- | --- | --- |
|  | N | % of all girls | N | % of all boys |  |
| EHCP only | 21710 | 2% | 62700 | 4% | 74% |
| SEN support only | 334880 | 24% | 541520 | 38% | 62% |
| CLA only | 8940 | 1% | 4320 | k | 33% |
| EHCP & CLA | 3500 | k | 9160 | 1% | 72% |
| SEN support & CLA | 14030 | 1% | 15300 | 1% | 52% |
| None | 985080 | 72% | 806110 | 56% | 45% |

*'k' is used when a result that is not zero would appear as zero due to rounding*
*CLA: children looked after, EHCP: education, health and care plan; SEN: Special Educational Needs*

**Appendix Table 2.** Admission rates (per 100 PY) aged 11-23 years old by gender and level of statutory provision.

|  |  | Planned |  |  | Unplanned |  |  | Mental-health related |  |  | Pregnancy-related |  |  |
| --- | --- | --- | --- | --- | --- | --- | --- | --- | --- | --- | --- | --- | --- |
|  |  | Rate | LCI | UCI | Rate | LCI | UCI | Rate | LCI | UCI | Rate | LCI | UCI |
| Girls | total | 4.83 | 4.82 | 4.84 | 5.35 | 5.34 | 5.36 | 2.56 | 2.55 | 2.57 | 4.00 | 3.99 | 4.01 |
|  | Ever CLA | 7.60 | 7.50 | 7.71 | 18.24 | 18.08 | 18.40 | 12.60 | 12.47 | 12.74 | 14.05 | 13.91 | 14.19 |
|  | ANY EHCP | 21.57 | 21.39 | 21.75 | 12.09 | 11.96 | 12.22 | 4.03 | 3.96 | 4.11 | 3.48 | 3.41 | 3.56 |
|  | ANY SEN support | 5.43 | 5.41 | 5.45 | 6.81 | 6.78 | 6.83 | 3.46 | 3.44 | 3.48 | 6.74 | 6.71 | 6.77 |
|  | None | 4.20 | 4.19 | 4.21 | 4.55 | 4.54 | 4.56 | 2.11 | 2.10 | 2.12 | 2.96 | 2.95 | 2.97 |
|  | only EHCP | 21.11 | 20.92 | 21.30 | 10.52 | 10.39 | 10.66 | 3.31 | 3.24 | 3.39 | 3.26 | 3.18 | 3.33 |
|  | only SEN support | 5.44 | 5.41 | 5.46 | 6.36 | 6.33 | 6.38 | 3.06 | 3.04 | 3.07 | 6.36 | 6.33 | 6.38 |
|  | only CLA | 4.77 | 4.63 | 4.91 | 17.68 | 17.41 | 17.95 | 13.35 | 13.11 | 13.58 | 14.54 | 14.30 | 14.79 |
|  | EHCP & CLA | 24.44 | 23.94 | 24.96 | 21.87 | 21.39 | 22.36 | 8.53 | 8.23 | 8.83 | 4.90 | 4.68 | 5.13 |
|  | SEN support & CLA | 5.22 | 5.10 | 5.34 | 17.70 | 17.48 | 17.92 | 13.15 | 12.96 | 13.34 | 16.02 | 15.81 | 16.22 |
| Boys | Total | 4.50 | 4.49 | 4.51 | 3.92 | 3.91 | 3.93 | 1.28 | 1.28 | 1.29 |  |  |  |
|  | Ever CLA | 8.40 | 8.30 | 8.51 | 10.63 | 10.52 | 10.75 | 5.34 | 5.26 | 5.42 |  |  |  |
|  | ANY EHCP | 13.07 | 12.99 | 13.16 | 7.24 | 7.18 | 7.30 | 2.42 | 2.38 | 2.45 |  |  |  |
|  | ANY SEN support | 4.48 | 4.46 | 4.49 | 4.47 | 4.45 | 4.48 | 1.62 | 1.61 | 1.63 |  |  |  |
|  | None | 3.77 | 3.75 | 3.78 | 3.22 | 3.21 | 3.23 | 0.94 | 0.93 | 0.94 |  |  |  |
|  | only EHCP | 12.39 | 12.30 | 12.47 | 6.24 | 6.18 | 6.30 | 2.00 | 1.96 | 2.03 |  |  |  |
|  | only SEN support | 4.48 | 4.47 | 4.50 | 4.33 | 4.32 | 4.35 | 1.51 | 1.50 | 1.52 |  |  |  |
|  | only CLA | 3.36 | 3.19 | 3.53 | 8.24 | 7.98 | 8.51 | 4.82 | 4.62 | 5.02 |  |  |  |
|  | EHCP & CLA | 17.80 | 17.53 | 18.07 | 14.13 | 13.89 | 14.37 | 5.29 | 5.15 | 5.44 |  |  |  |
|  | SEN support & CLA | 4.19 | 4.09 | 4.29 | 9.21 | 9.06 | 9.36 | 5.52 | 5.40 | 5.63 |  |  |  |

**Appendix Table 3.** proportion of pupils with at least one hospital admission during the follow-up period by exposure group and gender.

|  |  | Planned | Unplanned | Mental Health related | Pregnancy-related |
| --- | --- | --- | --- | --- | --- |
| **Girls** | EHCP only | 8,860 (41%) | 7,320 (34%) | 3,460 (16%) | 2,200 (10%) |
|  | SEN support only | 82,130 (25%) | 98,200 (29%) | 56,310 (17%) | 71,160 (21%) |
|  | CLA only | 2,220 (25%) | 4,550 (51%) | 3,690 (41%) | 3,530 (39%) |
|  | EHCP & CLA | 1,600 (46%) | 1,670 (48%) | 890 (25%) | 520 (15%) |
|  | SEN support & CLA | 3,580 (26%) | 7,050 (50%) | 5,640 (40%) | 6,010 (43%) |
|  | None | 217,650 (22%) | 238,240 (24%) | 126,720 (13%) | 112,120 (11%) |
| **Boys** | EHCP only | 19,250 (31%) | 17,700 (28%) | 7,790 (12%) |  |
|  | SEN support only | 117,190 (22%) | 142,530 (26%) | 58,080 (11%) |  |
|  | CLA only | 860 (20%) | 1,670 (39%) | 1,040 (24%) |  |
|  | EHCP & CLA | 3,590 (39%) | 3,900 (43%) | 2,190 (24%) |  |
|  | SEN support & CLA | 3,300 (22%) | 6,240 (41%) | 4,100 (27%) |  |
|  | None | 163,180 (20%) | 177,550 (22%) | 60,370 (7%) |  |

## Funding / Acknowledgements

This study is funded by the National Institute for Health and Care Research (NIHR) through the Children and Families Policy Research Unit (NIHR206114). The views expressed are those of the authors and not necessarily those of the NIHR or the Department of Health and Social Care. Research at UCL Great Ormond Street Institute of Child Health is supported by the NIHR Great Ormond Street Hospital Biomedical Research Centre. RB is supported by UK Research and Innovation [MR/Y030788/1] as part of Population Health Improvement UK (PHI-UK), a national research network that seeks to transform health and reduce inequalities through change at the population level.

## Data Availability

ECHILD data is available to external researchers and accessible via ONS Secure Research Service, as part of the ECHILD database. Interested researchers can apply to the ECHILD Data Access Committee. For more information on ECHILD and application process, please contact.

https://www.echild.ac.uk

